# Ankle proprioception in children with cerebral palsy

**DOI:** 10.1101/2023.04.26.23289148

**Authors:** Elizabeth Boyer, Qiyin Huang, Stacy Ngwesse, Jennifer Nelson, Jinseok Oh, Jürgen Konczak

**Affiliations:** Gillette Children’s – Center for Gait and Motion Analysis Saint Paul, Minnesota, USA; University of Minnesota - Twin Cities, Department of Orthopedic Surgery, Minneapolis, Minnesota, USA; University of Minnesota - Twin Cities, Human Sensorimotor Control Laboratory of School of Kinesiology, Minneapolis, Minnesota, USA; Gillette Children’s – Department of Research, Saint Paul, Minnesota, USA

**Keywords:** balance, human, position sense, somatosensory, kinesthesia

## Abstract

1.

**Purpose:** There is no established clinical standard to evaluate ankle proprioception in children with cerebral palsy (CP). This study compared ankle position sense of children with CP to age-matched children typically developing (TD).

**Methods:** Children aged 6-17 years participated (15 CP, 58 TD). Using a custom-built device, the ankle was passively rotated to 2 positions for 25 trials. Using a psychophysical forced-choice paradigm, participants indicated which position was more plantarflexed. A psychometric function was fitted to the response data to determine the just noticeable difference (JND) threshold and the associated uncertainty (random error) for ankle position sense.

**Results:** Median JND thresholds for the CP group were elevated (CP: 4.3°, TD: 3.0°). Three children with CP exceeded the 95^th^ percentile of TD. No differences in random error were found.

**Conclusion:** This method assessed ankle proprioception relative to norm data and identified position sense impairments in children with CP. Using this method can provide data on proprioceptive status in CP, augmenting the assessment of motor impairment.

## 2. INTRODUCTION

Proprioceptive impairment likely contributes to the high rate of falls reported in children with cerebral palsy (CP) [1]. However, proprioceptive status is almost never assessed clinically, and no comprehensive data exist on the breadth and severity of proprioceptive dysfunction in CP. Previous research examining lower extremity proprioceptive function in CP applied joint position matching or movement detection methods at various joints [2–11]. Yet, there are limitations to these approaches. For example, the movement detection method is commonly used in clinical sensory tests (e.g., Nottingham Sensory Assessment or Erasmus MC modified Nottingham Sensory Assessment; Rivermead Assessment of Somatosensory Performance) in which participants, with eyes closed, identify if or in which direction a limb was moved. The proportion of correct responses is summed over 3-6 trials. While quick, this method cannot quantify the extent of the proprioceptive deficit since it yields only dichotomous classification. Joint position matching methods address some of the above shortcomings but often they require participants to actively move their limb. Consequently, motor impairments may confound the proprioceptive (somatosensory) function one aims to measure [2–6,8,9,11,12]. Only Damiano et al. [2] accounted for this motor impairment by subtracting out the joint position matching error during a vision condition compared to a no-vision condition. Another problem with many existing methods is the lack of age-matched data of for peers typically developing (TD), making it difficult to contextualize proprioceptive impairment among individuals with CP [3,4,6,9,10]. Lastly, none of the previous CP proprioception studies used the *method of constant stimulus*, which is considered the most accurate of the 3 psychophysical methods to quantify proprioceptive acuity [13,14]. Acuity reflects the sharpness of a sense, that is, the ability to discriminate between small stimuli.

This study utilized the *method of constant stimulus*, in which participants were repeatedly presented pairs of joint positions in a 2-alternative forced-choice paradigm to determine their smallest perceived angular difference. This method affords the necessary sensitivity and accuracy lacking in the available clinical tests. Importantly, it provides information about 2 aspects of position sense acuity - the *bias* or systematic error in form of the just noticeable difference (JND) threshold, and *precision* or random error in form of the uncertainty area (UA). Here, the UA corresponds to the variability in making repeated judgements about ankle positions. The method requires an attentive participant, short-term memory to compare the pairs of stimuli, and more trials than the other two methods. The purpose of this proof-of-concept study was to document that the proposed method can be used in children with CP who can walk and is able to identify children with abnormal ankle position sense when compared to children TD. Based on moderate to large effect sizes for proprioceptive impairments observed in other studies [2,8,11,12], it was hypothesized that children with CP would collectively show evidence of impaired ankle position sense compared to a cohort TD.

## 3. METHODS

### 3.1 Participants

A power analysis using an effect size estimated from between group differences observed in Zarkou et al. informed the target sample size of children with CP [12]. For power = 0.8, alpha = 0.05, and Cohen’s d ≈ 0.96, a minimum of 15 participants per group were necessary to detect a statistically significant difference in ankle position sense acuity. Fifteen children with CP (*M* age: 11 years 10 months; *SD*: ± 2 years 10 months; range: 6 years 8 months – 15 years 9 months; 6 males, 9 females] and 58 children TD [*M* age: 12 years 3 months; *SD*: ± 3 years 2 months; range: 7 –17 years; 34 males, 24 females] were able to follow the instructions and complete the study (Table 1). All children with CP were recruited from patients undergoing or had recently undergone a clinical gait analysis at Gillette Children’s. All but four of them completed the proprioception test immediately after a 2.5-hour clinical gait analysis. Inclusion criteria were a diagnosis of CP, Gross Motor Function Classification System (GMFCS) level I-III, able to comprehend English, and expected to be able to follow instructions. Exclusion criteria were any surgery in the past 9 months or botulinum toxin injection in the past 3 months. All children TD were recruited and tested at the 2019 Minnesota State Fair. Inclusion criteria were 1) no reported history of central or peripheral nervous system disorder, 2) no current injury to the lower limbs, and 3) no exposure to chemotherapy which could have affected somatosensory and motor function. Before testing, children TD completed the footedness questionnaire to determine the dominant foot to be tested [15]. Appropriate written parental consent and child assent were obtained prior to data collection. The studies were approved by the University of Minnesota Institutional Review Board.

**Table 1.** Participant Demographics.

|  | **n** | **Age (y),**<br>mean<br>(SD) [range] | **Male**<br>**sex,**<br>n (%) | **GMFCS**<br>**level,**<br>n (%) | **Topography,**<br>n (%) | **Tone type,**<br>n (%) | **Prior Treatment,**<br>n (%) |
| --- | --- | --- | --- | --- | --- | --- | --- |
| Typically developing | 58 | 12 (3) [7-17] | 24 (41) |  |  |  |  |
| Cerebral palsy | 15 | 11 y 10 mo<br>[6 y 8 mo -15 y 10 mo] | 11 (73) | I: 6 (40)<br>II: 7 (47)<br>III: 2 (13) | Hemi: 6 (40)<br>Di: 7 (47)<br>Tri: 2 (13) | Spastic: 13 (87)<br>Mixed: 2 (13) | Orthopedic: 4 (27)<br>Selective dorsal rhizotomy:<br>3 (20)<br>Tone: 9 (60) |

Tone treatments included: intrathecal baclofen pump, botulinum toxin, phenol.
Di: diplegic, GMFCS: gross motor function classification system level, Hemi: hemiplegic, mo: months, n: sample size, SD: standard deviation, Tri: triplegic, y: years

### 3.2 Apparatus

Data were collected using the manual Ankle Proprioceptive Acuity System for all participants (Figure 1A). Feasibility of this system for measuring human ankle position sense acuity had been established previously [16]. Moreover, intra- and inter-rater reliability concerns are negligible because the applied psychophysical method is not subject to experimenter bias. The experimenter does not rate a perceiver’s performance but only rotates the joint to the position indicated by the psi marginal adaptive algorithm. With respect to test-retest reliability, the only source of variability is the inherent variability of the responder’s perception of ankle position. Unpublished data from eight healthy adults tested on three consecutive days showed very low test-retest variability (standard error of measurement = 0.09°).

**Figure 1.**
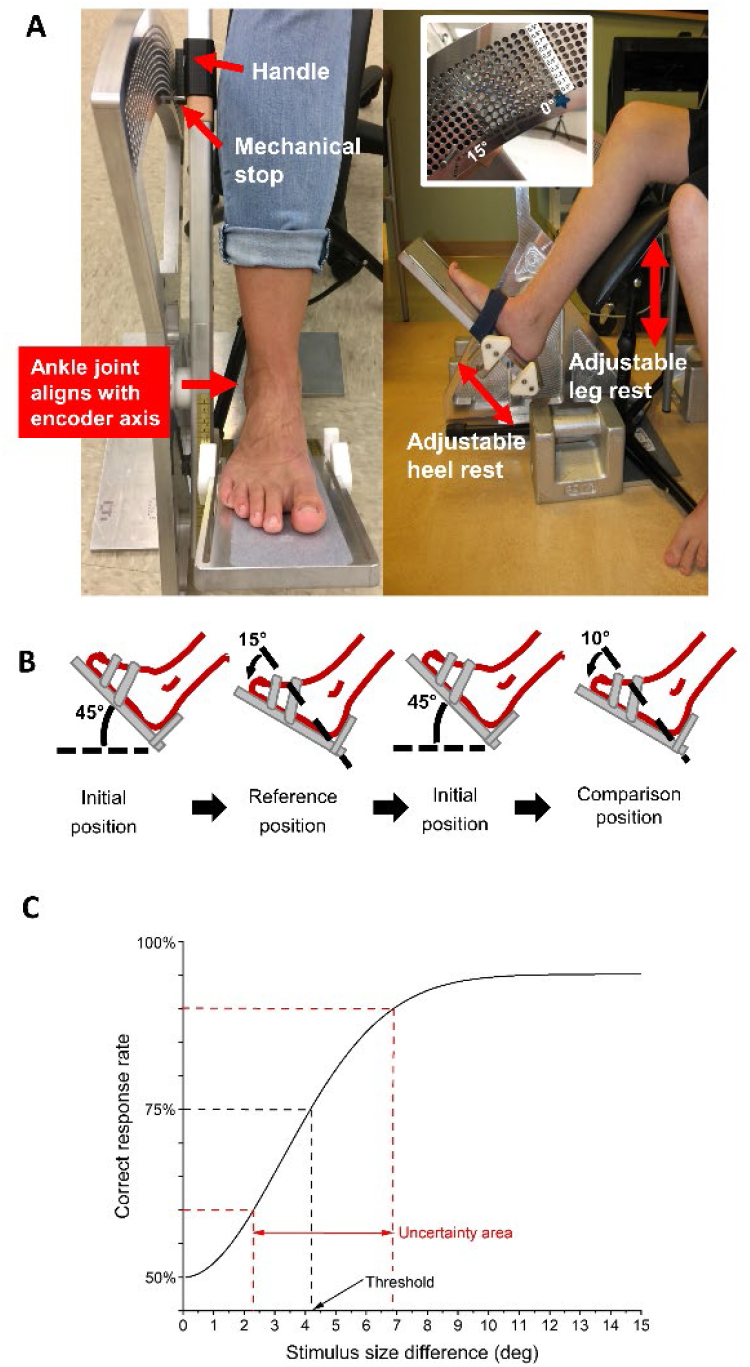
**A**. Front and side view of the manual Ankle Proprioceptive Acuity System. Rotating the handle by the experimenter rotates the ankle. Desired degree of rotation can be set by mechanical stops in the semicircular pegboard at 0.1° increments. The system components can be adjusted to the leg anthropometrics of the participant, so that the approximate ankle joint axis aligns with the axis of rotation of the device. **B**. Example of the time course of a single trial. All trials started at the neutral 45° position. Reference position was always at 15° plantarflexion. Here the researcher rotates the foot to the reference position, then returns the footrest to the initial position before rotating it to the comparison position (10° plantarflexion). Thus, the stimulus difference is 5°. The participant indicates which experienced position (1^st^ or 2^nd^) was closer to the floor. **C**. Example of a derived psychometric function. The JND threshold corresponds to the difference between reference and comparison position at the 75% correct response rate. The uncertainty area is defined as the distance between the stimulus size differences at the 60^th^ and 90^th^ percentiles.

The system allows for manual plantarflexion/dorsiflexion of the tested ankle to distinct ankle positions with a pegboard position resolution of 0.1°. Angular position and velocity were recorded by a U.S. Digital H6 Optical Encoder (resolution: 0.036°) embedded in the device and aligned with the participant’s ankle joint axis (sampling frequency 100 Hz).

### 3.3 Assessment procedure

Participants were barefoot and seated on a chair to perform the ankle position discrimination test on the more-involved (CP) or dominant ankle (TD). The footrest height and heel rest position were adjusted to align the ankle center, approximated as the lateral malleolus, with the device axis of rotation. The tested foot, stabilized by a strap, sat on the footrest with the participant’s ankle at approximately 90° relative to the shank. Participants wore vision occluding glasses to remove possible visual cues of ankle position during testing.

One of three researchers slowly plantarflexed the participant’s foot from an initial neutral position (0° plantarflexion) to a reference (15° plantarflexion) or comparison (0-15° plantarflexion) position. The participant’s foot was held for approximately 2 seconds, then moved back to the initial neutral position before moving to the other reference or comparison position (Figure 1B). Participants were asked to verbally indicate in which of the 2 positions (first or second) their toes were closer to the floor. Participants were told to provide their best guess if they were unsure. They were allowed to repeat a trial if they were not paying attention. Before the testing, there were at least 3 practice trials with or without vision occluding glasses to help participants acclimate to the device and testing procedure. The practice trials began with a large difference between the reference and comparison position (approximately 10°), to assure that participants were able to discriminate and to understand the testing scheme.

During actual testing, a participant’s verbal response (incorrect/correct) in the previous trial was used as input for an adaptive psi-marginal algorithm to determine the comparison position for the next trial within the allowable 15° stimulus range [17]. A total of 25 trials were performed. A break was given after 10 trials or when participants requested. Instructions were reiterated when necessary. The complete protocol including practice and 25 test trials took 15-30 minutes to complete.

### 3.4 Psi marginal adaptive algorithm

The psi marginal adaptive algorithm was used to update which comparison ankle position to present the participant for each test trial [17]. The four parameters for this algorithm include the threshold (alpha), slope (beta), upper asymptote (i.e., lapse rate; lambda), and lower asymptote (i.e., guess rate; gamma). The threshold was of primary interest and was the stimulus intensity (difference between the reference and comparison ankle positions, in °) at which a participant would discriminate two different ankle positions at 75% accuracy. The threshold was set to be searched within the range of 0-15° of plantarflexion with an increment of 0.1°. The slope measures the variability in perceptual judgments during an assessment. The slope was set to be searched within the range of -1.2 to 1.2. Lapse rate represents the proportion of trials the participant incorrectly answered because of inattentiveness for stimuli that the participant is truly capable of differentiating when vigilant. Lapse rate was set to be searched within the range of 0 to 0.1. Guess rate for this two-alternative forced choice task was 0.5, meaning that participants could have guessed correctly 50% of the time. Wait time was set to 4, which represents the number of trials after the maximum stimulus intensity was presented (15° in this study) in which lapse rate is assumed fixed (0.1 in this study) so if a wrong answer occurred during this wait time, lapse rate was not updated in the posterior but rather stayed fixed.

### 3.5 Psychometric Function Fitting

A logistic Weibull function (i.e., Gumbel function) was fitted to the stimulus difference-response data (i.e., the difference in the reference and comparison ankle angular positions and the incorrect/correct verbal response data for each participant) [18]. Based on the fitted function (Figure 1C), 2 components of ankle position sense acuity were calculated: bias (systematic error) and precision (random error). *Bias* is measured as the JND threshold, the stimulus difference between the reference and comparison angular positions (in degrees) at the 75% probability of correct response of the function [13,19]. Smaller JND thresholds (or less bias) represent higher ankle position sense acuity, which implies that the person can discriminate between smaller differences in ankle position. *Precision* reflects how consistently a participant responds across all trials, represented by the uncertainty area (UA) of the fitted function. UA was calculated as the range of the stimuli (in degrees) between the 60% and 90% probabilities of a correct response [19]. A smaller UA value represents a higher ankle position sense precision, that is, the participant was more certain in his/her responses. The function fit allowed a range of UA ≤25°.

### 3.6 Statistical analysis

The statistical analysis was based on custom-written code in R (version 4.1.2). Shapiro-Wilk tests were performed to determine the normality of the outcome variables. Given that all variables were non-normal, a subsequent non-parametric Wilcoxon rank sum tests was applied to determine group differences (TD vs. CP) for JND threshold and UA. Statistical significance was defined as p < 0.05.

## 4. RESULTS

All individual JND threshold data of children with CP were compared to the age-appropriate median and quartiles from the cohort TD (Figure 2). Most children with CP (9/15; 60%, standard error: 13%) exhibited JND thresholds within the third and fourth quartiles of the cohort TD, with 3 children (20%, standard error: 10%) exhibiting thresholds above the 95^th^ percentile. There was no clear indication that JND threshold was related to a child’s GMFCS level. With respect to discerning differences at the group level (CP vs. TD), the Wilcoxon rank sum test showed a significant difference (W = 243.5, p = 0.009) between the JND threshold in children TD compared to children with CP. The median JND threshold was 3.0° (range: 0.9 - 7.4°) compared to 4.3° (range: 1.3 - 12.3°), respectively (see Figure 3).

**Figure 2.**
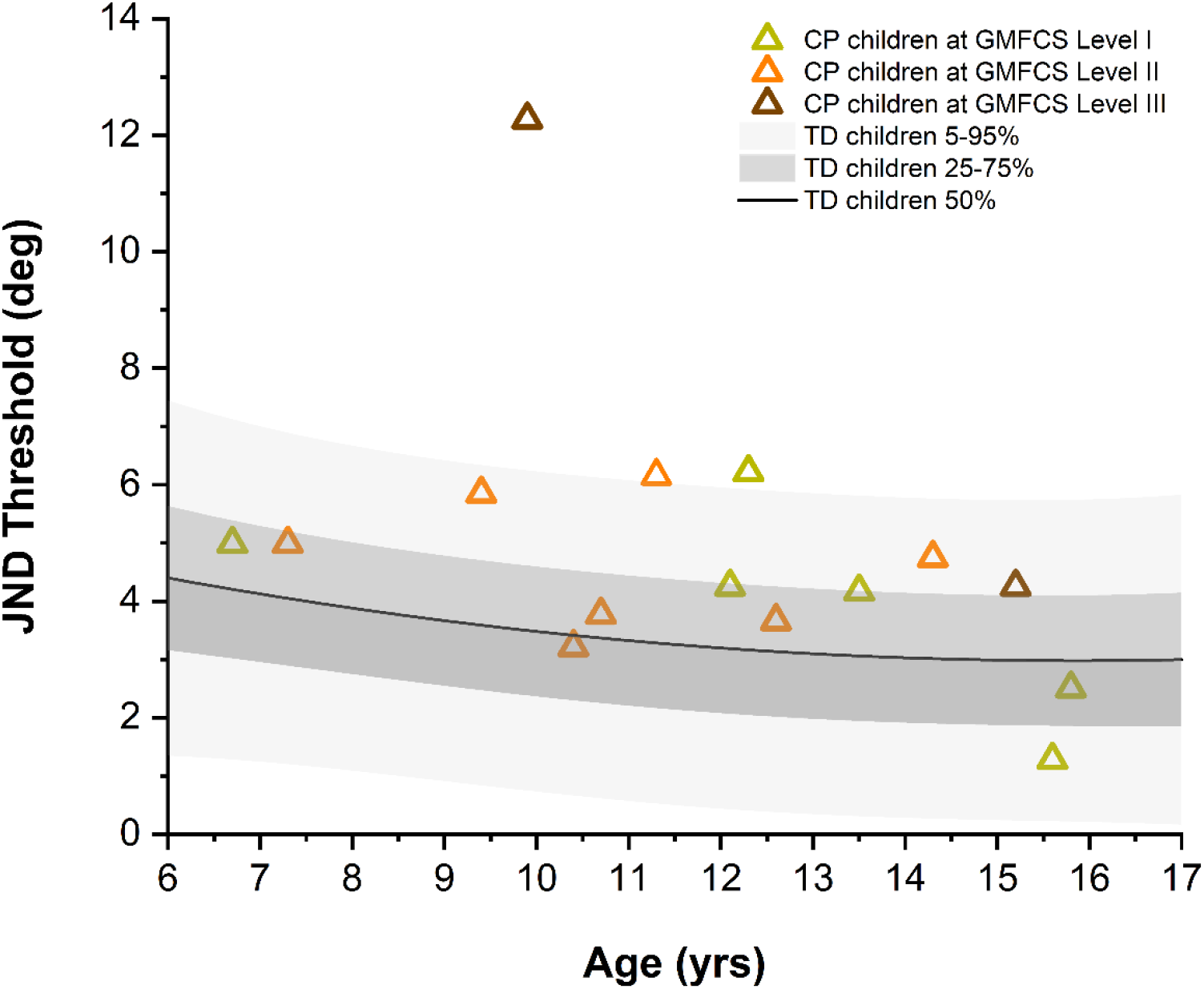
JND thresholds of all CP participants relative to chronological age of the cohort TD. Data of those TD were fitted with a 2^nd^ order polynomial function. Black line represents the median, dark and light gray bands represent the distribution between the 25-75^th^ and the 5-95^th^ percentiles, respectively.

**Figure 3.**
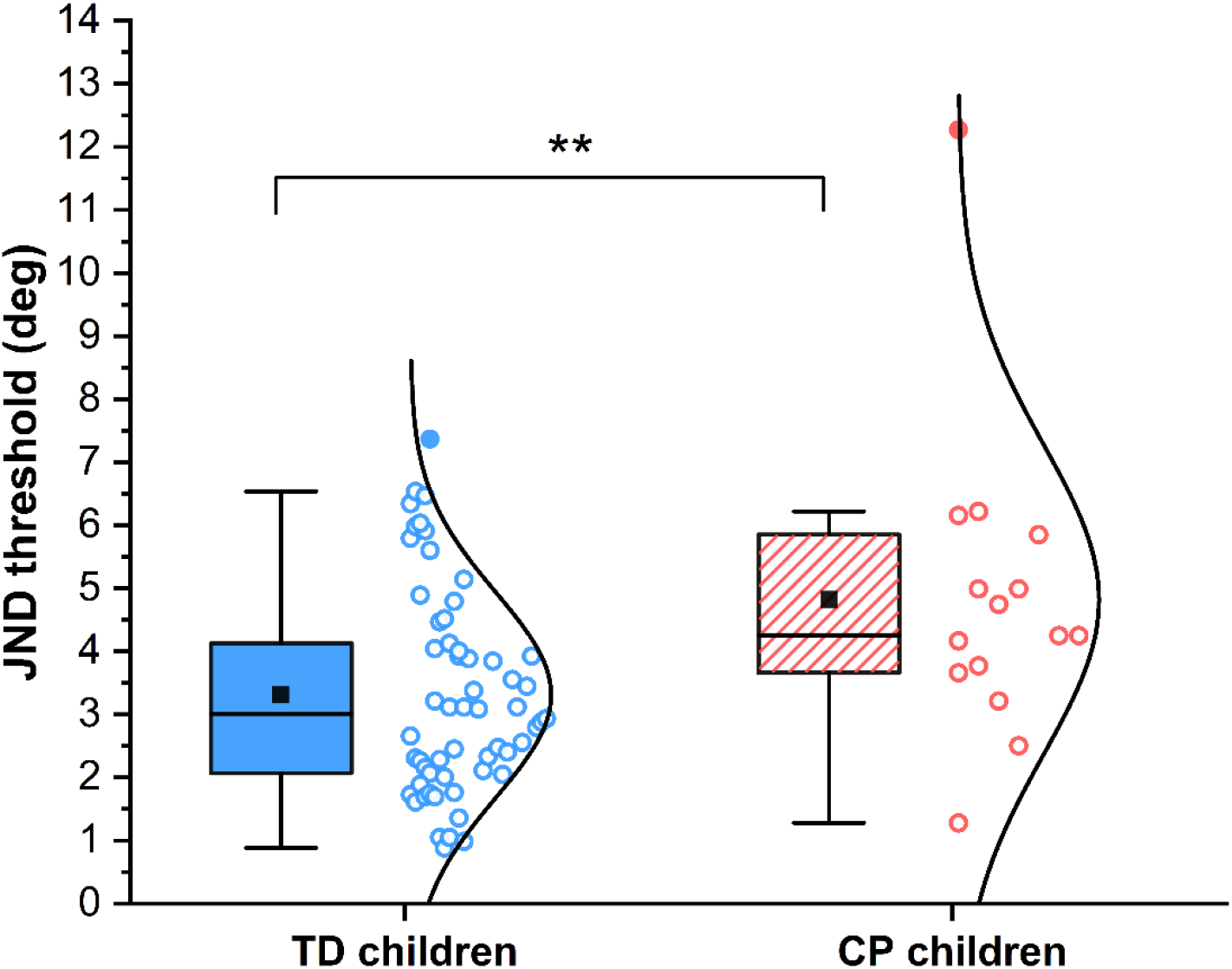
Bias of ankle position sense as measured by the JND threshold. The lower and upper limit of each box represents the 25^th^ and 75^th^ percentile, the whiskers the 5^th^ and 95^th^ percentile. Circle symbols represent individual participants, with filled circles representing participants that fell outside 1.5 × the interquartile range of their respective group. ** indicates statistically significant between-group differences.

The UA (precision or random error) of ankle position sense acuity was unaltered in these children with CP with respect to children TD (Figure 4). The median UA was 4.6° (range: 0.6 - 22°) in the group with CP and 3.4° (range: 0.4 - 17.6°; W = 353, p = 0.27) in the group TD.

**Figure 4.**
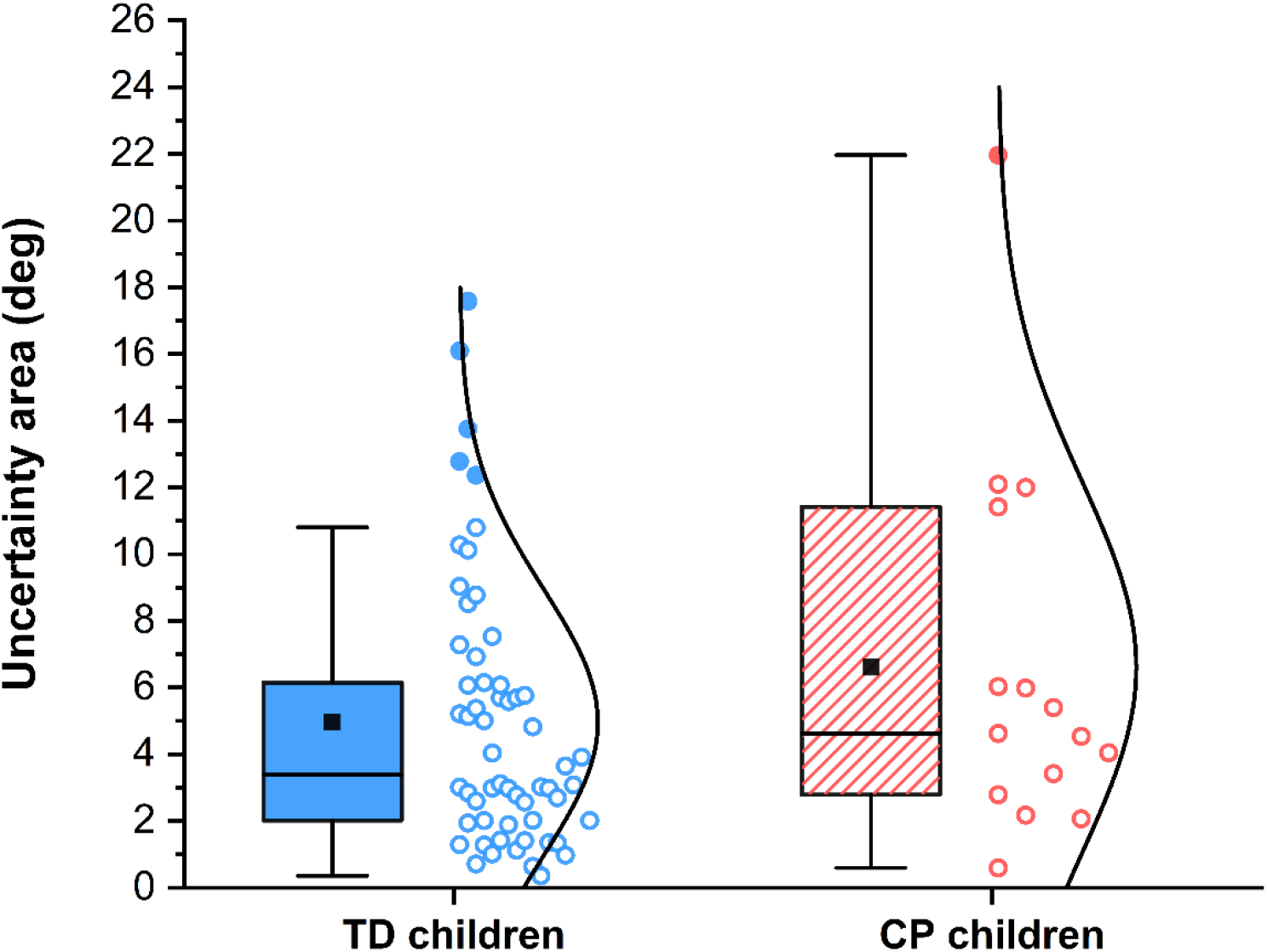
Precision of ankle position sense as measured by the uncertainty area (UA). The lower and upper limit of each box represents the 25^th^ and 75^th^ percentile, the whiskers the 5^th^ and 95^th^ percentile. Circle symbols represent individual participants, with filled circles representing participants that fell outside 1.5 × the interquartile range of their respective group.

## 5. DISCUSSION

The goal of this study was to objectively quantify ankle position sense acuity (bias and precision) in a pediatric CP sample. The results support the hypothesis that the CP group tends to have impaired ankle position sense. When considering these markers of proprioceptive function, it is important to note that both groups presented with higher thresholds than healthy young adults (2.4°) [16], indicating that ankle position sense acuity continues to develop until young adulthood [20].

The results suggest that impaired ankle position sense can be a clinical feature of children with CP in GMFCS levels I-III since 20% exhibited JND thresholds above the 95^th^ percentile of TD. This also means that approximately 80% may present with ankle position sense JND thresholds within the range of children TD, though most of the CP group were distributed above the median. In fact, the median JND threshold of the CP group was 43% higher than the group TD. This study applied a paradigm where the ankle was passively rotated to exclude any possible confounds due to impaired motor function in CP. The between-group differences align with results reported by Zarkou et al. who used an active plantarflexion joint position matching task [12]. Both studies included individuals in GMFCS level I-III. Currently, there are no data to indicate that the degree of proprioceptive dysfunction observed in CP is correlated with symptom severity as determined by GMFCS level. However, it is known that the initial brain injury experienced by children with CP may damage ascending somatosensory tracts and their projections in addition to the descending motor pathways [21–23]. This finding provides a neurophysiological rationale for expecting proprioceptive deficits in CP, observed here as between-group differences.

There was no evidence that precision, the random error in making repeated judgements about ankle positions, was systematically affected in the group with CP. It is known from other studies of upper limb proprioception in children TD that proprioceptive bias already reaches adult levels in early childhood, but proprioceptive precision continues to improve until late adolescence [24]. In this study, the precision of ankle position sense in children with CP fell within the range of the age-matched cohort TD. If confirmed in a larger sample, this implies proprioceptive dysfunction in CP is characterized by a shift in proprioceptive bias not random error. This contrasts with reports on children with developmental coordination disorders. As a group, those children do not exhibit elevated upper extremity JND thresholds but have higher values of random error when making judgements about wrist and elbow positions [25].

Implementing clinical evaluations of proprioceptive function and expanding research in this area will help the field to establish the prevalence and magnitude of proprioceptive deficits in CP and to relate such somatosensory impairment to the observable motor problems. Having access to this knowledge has implications for prognosis and rehabilitation. Someone with widespread proprioceptive impairments may have a poorer gross motor prognosis than someone with intact proprioceptive function but profound weakness, which is more amenable to resistance training. However, there is evidence that proprioception or somatosensory training can improve proprioceptive and motor function in other populations, as well as the upper extremity in CP [26,27]. Future studies should explore if that effect replicates in the lower extremity of the CP population.

### 5.1 Study limitations

There are several limitations to this study that need to be considered. First, this proof-of-concept study only examined a relatively small group of children with CP, which focused on high functioning children at GMFCS levels I-III. Currently, there are limited or no data on children in GMFCS levels IV-V. Yet, it would be imperative to obtain such data to discern how proprioceptive dysfunction involving the ankle contributes to deficits in gross motor ability. Second, like all psychophysical testing procedures, the method applied here, requires that the examinee has the cognitive ability to comprehend the task and stay focused and attentive during testing. This limits the applicability of the test to school-age children and approximately half of individuals with CP who do not have an intellectual disability [28]. Variability of maintaining attention, especially in younger children, was evident in this study. Only 29% (2/7) of the children with CP younger than 9 years old successfully completed testing compared to 62% (13/21) of children 9-15 years old. Such an age dependency to obtain valid data has also been observed by others performing sensory testing with children [5].

Finally, high lapse rates can spuriously increase JND thresholds [18,29,30]. While the participants’ lapse rate in this study was not objectively measured, it was assumed that it ranged from 0-30% as others have observed [29,30]. If true lapse rate (denoted as lambda in the psychophysical function) was greater than the estimated lambda from the adaptive psi-marginal algorithm (restricted to ≤10%), JND thresholds presented here would be overestimated, indicating they have poorer acuity than their true ability [18]. However, there is a practical trade-off between more precise estimates of JND threshold and UA by collecting more trials and increased likelihood of attentional lapses with longer testing.

## 6. CONCLUSIONS

This study provides initial evidence that higher functioning children with CP can present with impaired ankle position sense acuity, but such proprioceptive impairment may not be widespread. The outcome measures herein most closely represent “pure” proprioceptive function as it does not rely on active movement which would make it difficult to distinguish between sensory and motor impairment in CP. This research begins to address the knowledge gap around prevalence and severity of lower extremity proprioceptive impairment in individuals with CP. It outlines an approach to more systematically investigate how proprioceptive impairment affects balance control, whether it is associated with higher fall frequency, and how it impacts the motor learning ability of these children.

## Data Availability

All data produced in the present study are available upon reasonable request to the authors.

## 7. ACKNOWLEDGEMENTS

The authors extend their gratitude to all participants for volunteering for this study and Dr. Holst-Wolf for help with data collection and training.

This work was supported by the Endowed Fund for Research in Cerebral Palsy Treatment and Gait and Motion Outcomes Fund of Gillette Children’s and research funds by Dr. Konczak. The device for assessing ankle proprioception was designed by the Human Sensorimotor Control Laboratory of University of Minnesota.

## 9. ETHICAL CONSIDERATIONS

These studies (STUDY00009461 and STUDY00003446) were approved by the University of Minnesota Institutional Review Board on May 15^th^, 2020 and May 3^rd^, 2019 respectively. Informed consent was obtained for all participants in both studies.

